# Impact of subgroup classificuation accuracy on detecting heterogeneous treatment effects in *Staphylococcus aureus* bacteraemia: A simulation study

**DOI:** 10.64898/2026.07.17.26357924

**Authors:** Fergus Hamilton, Sean Ong, Maaike C Swets, Clark D. Russell, Jonathan Underwood

## Abstract

Staphylococcus aureus bacteraemia (SAB) is clinically heterogeneous. Potential heterogeneous treatment effects (HTE) have recently been identified through analysis of patient subgroups, identified using clinical variables. However, the impact of misclassifying patients into these groups is unclear, and strategies to improve HTE detection remain uncertain.

**Methods:** We performed a simulation study using data from selected randomised trials and observational studies in SAB. We assessed the impact of varying classification accuracy (70%-100%) on i) power, ii) type-I error, and iii) bias in post-hoc analyses of HTE. We then evaluated two strategies to improve performance: enrichment designs, in which only patients predicted to belong to a target subgroup are randomised, and the use of ordinal rather than binary outcomes.

**Results:** Even with perfect classification, post-hoc detection of heterogeneous treatment effects remained highly conditional on subgroup prevalence, baseline mortality, and effect size. One subgroup was detectable at moderate sample sizes; however, power was inadequate for all other subgroups even with sample sizes of 20,000.

Decreasing classification accuracy reduced power, increased type-I error, and introduced bias. Enrichment marginally improved power. Ordinal outcomes substantially improved performance when they matched the treatment-effect structure, but were worse when they did not.

**Conclusions:** Detecting HTE in SAB is challenging, but not uniformly infeasible. Feasibility depends on the interaction between subgroup frequency, baseline risk, classifier performance, and outcome choice. To advance stratified medicine in SAB, research should prioritize robust classifiers, outcome measures matched to the expected mechanism of treatment effect, and trial designs that acknowledge uncertainty in key parameters.

## Introduction

Staphylococcus aureus bacteraemia (SAB) is a common and serious infection associated with significant morbidity and mortality.^1,2^ A defining feature of SAB is its clinical heterogeneity, encompassing variations in patient characteristics (e.g., age, comorbidities), pathogen factors (e.g., methicillin-resistance), portal of entry of bacteraemia, and disease manifestations such as location of infection and metastatic foci.^3^ Despite this heterogeneity, clinical trials in SAB often treat it as a single entity, introducing the risk of failing to detect treatments which differentially benefit or harm subgroups of patients.^4,5^ In particular, trials investigating adjunctive or alternative therapies have frequently failed to show overall benefit compared to standard care, although recent data suggests antimicrobial choice affects outcomes.^5–7^

Recent efforts have focused on identifying clinically relevant subgroups of patients with SAB to enable better patient stratification for research and potentially personalised treatment. Using latent class analysis on routinely available data from observational and trial cohorts from the UK, Spain, the Netherlands, and the USA, investigators identified five distinct and reproducible subphenotypes (subphenotypes A-E),^8,9^ while other studies have taken similar approaches.^10,11^ For consistency with prior literature, these are referred to as subgroups henceforth throughout.

Importantly, these analyses suggested potential heterogeneous treatment effects^12^ (HTE) of adjunctive rifampicin across these subgroups. Notably, rifampicin appeared potentially harmful in subgroup B (nosocomial IV catheter-associated SAB; OR 18.8 for 84-day mortality), although the effect size is likely inflated given 84-day mortality was zero in the placebo group.^8,13^

The possibility of such HTE raises important questions for future trial design and for stratified medicine in SAB. In principle, participants in an RCT, or patients in clinical practice, could be stratified into subgroups and then offered treatments more likely to help them. This may be preferable to applying the same therapy across an unselected SAB population, particularly when the biological plausibility of benefit differs across subgroups. For example, in nosocomial catheter-associated bacteraemia, where catheter removal is the main intervention, the potential benefit of adjunctive antimicrobial strategies may be smaller than in patients with a higher burden of infection, such as those with endocarditis or native vertebral osteomyelitis.

If treatment effects truly differ between subgroups, accurately identifying these becomes paramount. However, any diagnostic test or classification algorithm used to assign patients to subgroups will inevitably have imperfect accuracy.

Misclassification can bias subgroup-specific treatment-effect estimates, inflate type I error, and reduce power to detect true HTE, potentially leading to misleading conclusions about which patients benefit from or are harmed by treatment.

Understanding the quantitative impact of misclassification is essential for interpreting subgroup analyses and designing efficient trials, including potential enrichment strategies and the use of ordinal outcomes. The choice of outcome measure may also affect both power and susceptibility to this bias. For example, odds ratios are commonly used to compare mortality between treatment groups, but they are a non-linear transformation of risk. Consequently, subgroups with different baseline mortality may have similar odds ratios but substantially different absolute treatment effects. This property - known as non-collapsibility - can make comparisons of odds ratios across low- and high-risk subgroups less intuitive, particularly when risks differ markedly.^14^

## Methods

To investigate the interplay between diagnostic accuracy, sample size, and statistical power in trials with substantial treatment-effect heterogeneity, we conducted a comprehensive simulation study using the aims, data generation, estimands, methods, and performance measures (ADEMP) framework.^15^ The study was designed to be directly relevant to clinical trials in SAB, with parameters for treatment effects, subgroup frequencies, and baseline risk taken from recent published literature on SAB subgroups.^8,9^ The simulation framework was implemented in R (version 4.5.1) using tidyverse-based scripted analyses. Full details of the simulation study are in the **Supplementary Methods**. We provide a brief overview here.

### Main analysis

In our main analysis we simulated a simple two-arm randomised trial with subgroup-specific treatment effects derived from analyses of the ARREST trial^5^Because subgroup-specific treatment effects estimated in ARREST can be affected by the ‘winner’s curse’we applied a 50% shrinkage on the log-odds scale to define the prespecified primary effects. For simplicity, we used the average mortality across both arms given the trial had no overall effect.^13^

The primary outputs of this simulation were type I error (significant effect in a null subgroup), power (significant effect in the correct direction), and bias (difference between estimated and true subgroup log-odds ratio).

### Supplementary analysis 1: Sensitivity to subgroup prevalence and mortality

Across cohorts in which SAB subgroups have been measured, both subgroup prevalence and within-subgroup mortality vary substantially. We therefore tested how sensitive the simulations were to these changes in prevalence and mortality while holding the subgroup treatment effect constant.

### Supplementary analysis 2: enrichment feasibility

We then evaluated enrichment designs under varying sensitivity/specificity to quantify effect dilution and screening burden (number needed to randomise and number needed to screen). This tests a scenario where a classifier could be applied at or just before randomisation, and only those in certain subgroups are enrolled.

### Supplementary analysis 3: ordinal outcome

Ordinal outcomes can provide greater precision when they capture clinically meaningful gradations of severity. In this analysis we compared binary and ordinal endpoints under the same subgroup-effect structure and misclassification settings to assess gains in precision and their relationship to bias. Specifically, we used a 6-point ordinal scale spanning death to recovery. We evaluated two scenarios: one with a proportional-odds effect across the full ordinal scale, and one with a non-proportional death-only effect in which treatment changes only death odds while the other non-death categories are rescaled proportionally. As a sensitivity analysis, we also evaluated the 5-point ordinal scale. The death-only scenario represents a clinically plausible pattern in which an intervention affects mortality but has little or no effect on less severe outcome levels; this pattern has been observed in infection trials and allows assessment of when ordinal analyses may dilute a clinically important mortality signal.

## Results

### Sample-size requirements for a two-arm RCT of subgroups

The statistical power to detect subgroup-specific treatment effects in a standard two-arm RCT, assuming perfect classification and the prespecified shrunk effects, was highly dependent on subgroup prevalence, baseline event rate, and effect size. The results, summarised in **Table 2** and **Figure 1**, show that HTE in subgroup B was detectable at realistic sample sizes, reaching essentially complete power by n=3,000. In contrast, all other subgroups remained underpowered even at n = 20,000. Fuller operating characteristics are provided in **Supplementary Table S1**.

**Figure 1:**
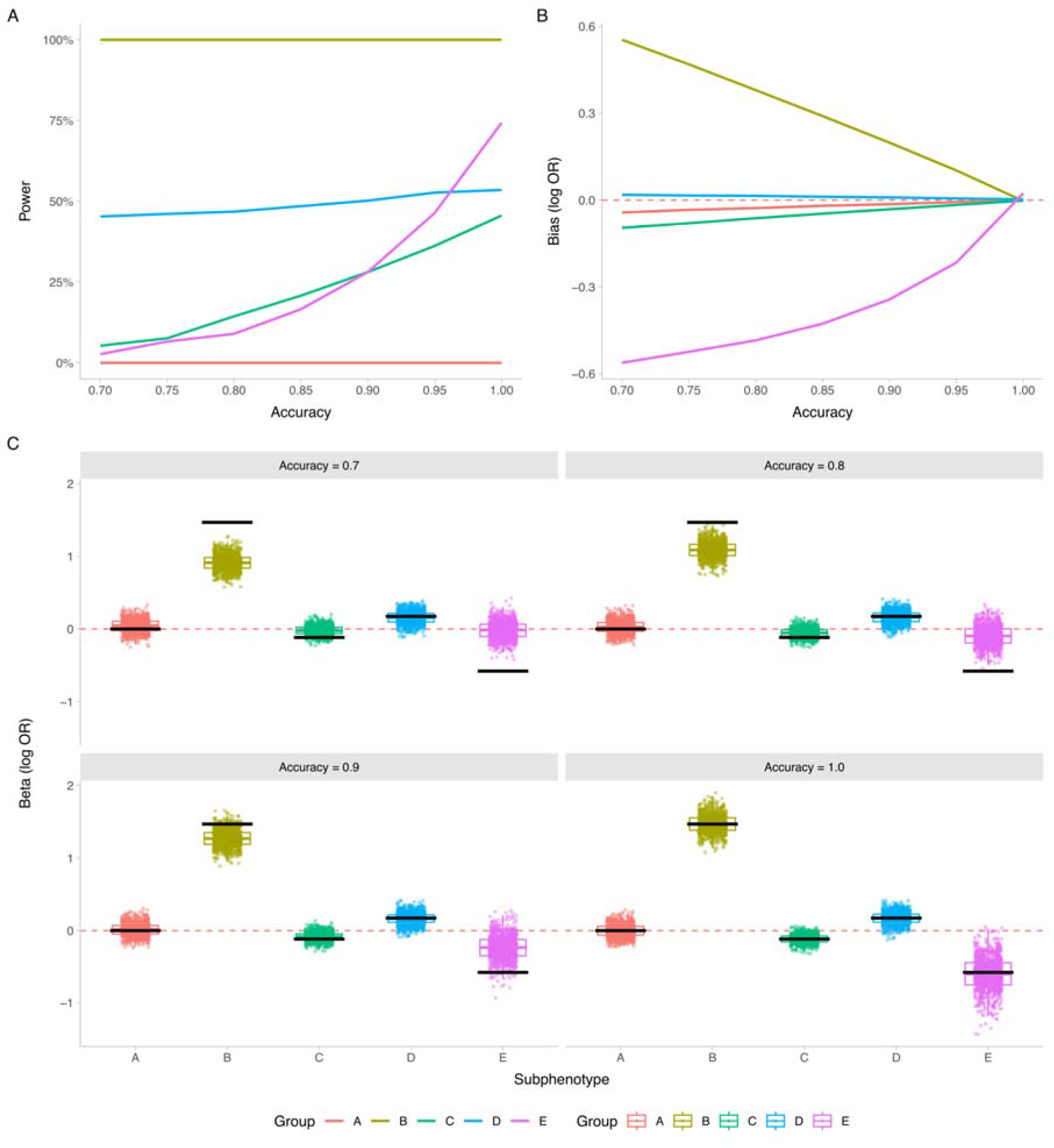
Impact of subgroup misclassification on subgroup-specific effect estimation. A) Power versus classification accuracy. B) Mean bias (true logOR minus estimated logOR) versus accuracy. C) Replicate-level subgroup estimates by accuracy; boxplots show median and IQR, whiskers represent 1.5 x IQR, and black horizontal bars indicate true subgroup logOR (84-day mortality, treatment versus control).

**Table 1:** Main estimation parameters, with the shrunk OR representing a 50% decrease on the log-odds scale.

| Subgroup | Frequency in ARREST placebo arm (%) | ARREST 84-day mortality (%) | Primary OR for 84-day mortality |
| --- | --- | --- | --- |
| A | 15.5 | 21.7 | 1.00 |
| B | 13.4 | 7.5 | 4.34 |
| C | 35.6 | 19.2 | 0.89 |
| D | 17.8 | 18.2 | 1.19 |
| E | 17.8 | 2.9 | 0.56 |

**Table 2:** Statistical power to detect a subgroup-specific effect at each sample size n. Power was estimated from 2,000 simulated replicates. Subgroup A has no treatment effect, so power is not the appropriate metric.

| Total trial size | Subgroup A (null) | Subgroup B | Subgroup C | Subgroup D | Subgroup E |
| --- | --- | --- | --- | --- | --- |
|  | A | B | C | D | E |
| 500 | - | 0.408 | 0.054 | 0.040 | 0.000 |
| 1,000 | - | 0.820 | 0.059 | 0.075 | 0.004 |
| 2,000 | - | 0.986 | 0.089 | 0.086 | 0.037 |
| 3,000 | - | 1.000 | 0.110 | 0.112 | 0.112 |
| 5,000 | - | 1.000 | 0.155 | 0.179 | 0.228 |
| 10,000 | - | 1.000 | 0.272 | 0.311 | 0.421 |
| 20,000 | - | 1.000 | 0.486 | 0.527 | 0.708 |

As expected in a traditional two-arm RCT, Type I error was well calibrated and bias was minimal.

We then investigated the impact of imperfect classification accuracy on power, Type I error, and bias in a deliberately large hypothetical trial of 20,000 participants. As shown in **Figure 1**, statistical power to detect true effects diminished substantially as classification accuracy decreased. For the null subgroup A, this increased Type I error, while for all subgroups effect estimates became biased. The degree of bias depended both on the amount of misclassification and on the composition of the groups contributing those misclassified patients. This dilution effect occurs because misclassification mixes patients from subgroups with different true effects, pulling the observed estimate towards the population average. The effect was particularly marked for less frequent subgroups with large effects: the strong effect in subgroup B (OR=4.3 after shrinkage) was substantially attenuated by even small amounts of misclassification. The three panels of Figure 1 show the full accuracy analysis.

### Results are highly conditional on baseline subgroup prevalence and mortality even with a perfect classifier

It is important to understand how conditional these estimates are on the cohort under study, because sample-size calculations are made before a trial starts and the extent to which subgroup prevalence and mortality vary across settings is uncertain. We therefore reran the analyses using data from several cohorts with differing subgroup prevalences and mortality, assuming perfect classification. For most subgroups, the required sample size varied substantially by setting, in some cases by as much as five-fold (**Figure 2**; **Supplementary Table S2**).

**Figure 2:**
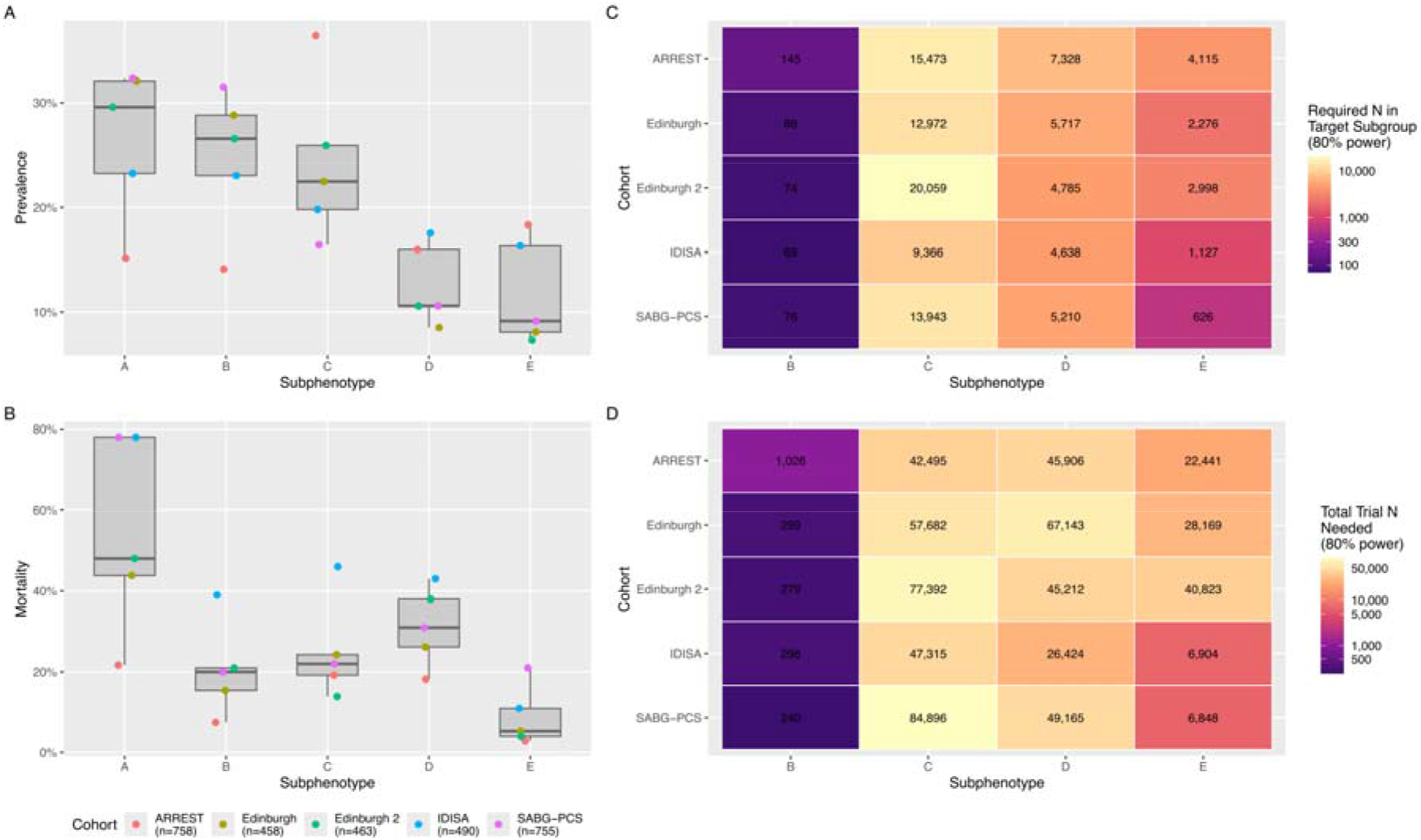
Effects of subgroup prevalence and mortality on sample-size requirements. A) Subgroup prevalence by cohort (points are cohorts; boxplots show median, IQR, and 1.5 x IQR whiskers). B) Subgroup-specific mortality by cohort. C) Heatmap of required N in target subgroup for 80% power. D) Heatmap of required total trial N for 80% power. Abbreviations: SABG-PCS, Staphylococcus aureus Bacteremia Group Prospective Cohort Study; IDISA, Improved Diagnostic Strategies in Staphylococcus aureus bacteremia study.

These findings show that even establishing an a priori sample-size calculation for HTE detection is difficult, because the planned trial setting is unlikely to mirror the subgroup distribution seen in other settings. Supporting this, the predicted total sample-size requirement differed meaningfully even between the two Edinburgh cohorts, reflecting relatively small but still important differences in subgroup prevalence and mortality.

### Enrichment strategies are unlikely to succeed when classifiers are poor for most subgroups

We then extended this by considering a hypothetical enrichment design in which patients with SAB are screened using an imperfect classifier and only those predicted to belong to a target subgroup are randomised prospectively.

We evaluated the impact of five hypothetical diagnostic tests ranging from mediocre to near-perfect accuracy on the number needed to screen (NNS) and number needed to randomise (NNR). For subgroup B, which combines a large treatment effect (OR 4.3 after shrinkage) with moderate frequency (13%), enrichment remained realistic: NNS ranged from about 1,100 with a near-perfect test (sensitivity and specificity 99%) to about 3,300 with a balanced test (sensitivity and specificity 80%), with corresponding NNR values of roughly 140 to 930 participants (**Figure 3**; **Supplementary Table S3**). Biased estimates were still present, as expected with imperfect classifiers. In contrast, for the other subgroups enrichment remained unattractive or infeasible even with excellent classifiers. Type I error in the null subgroup (A) remained close to nominal in the main simulation (n=20,000), ranging from 5.0% at 100% accuracy but rising modestly to 7.0% at 70% accuracy.

**Figure 3:**
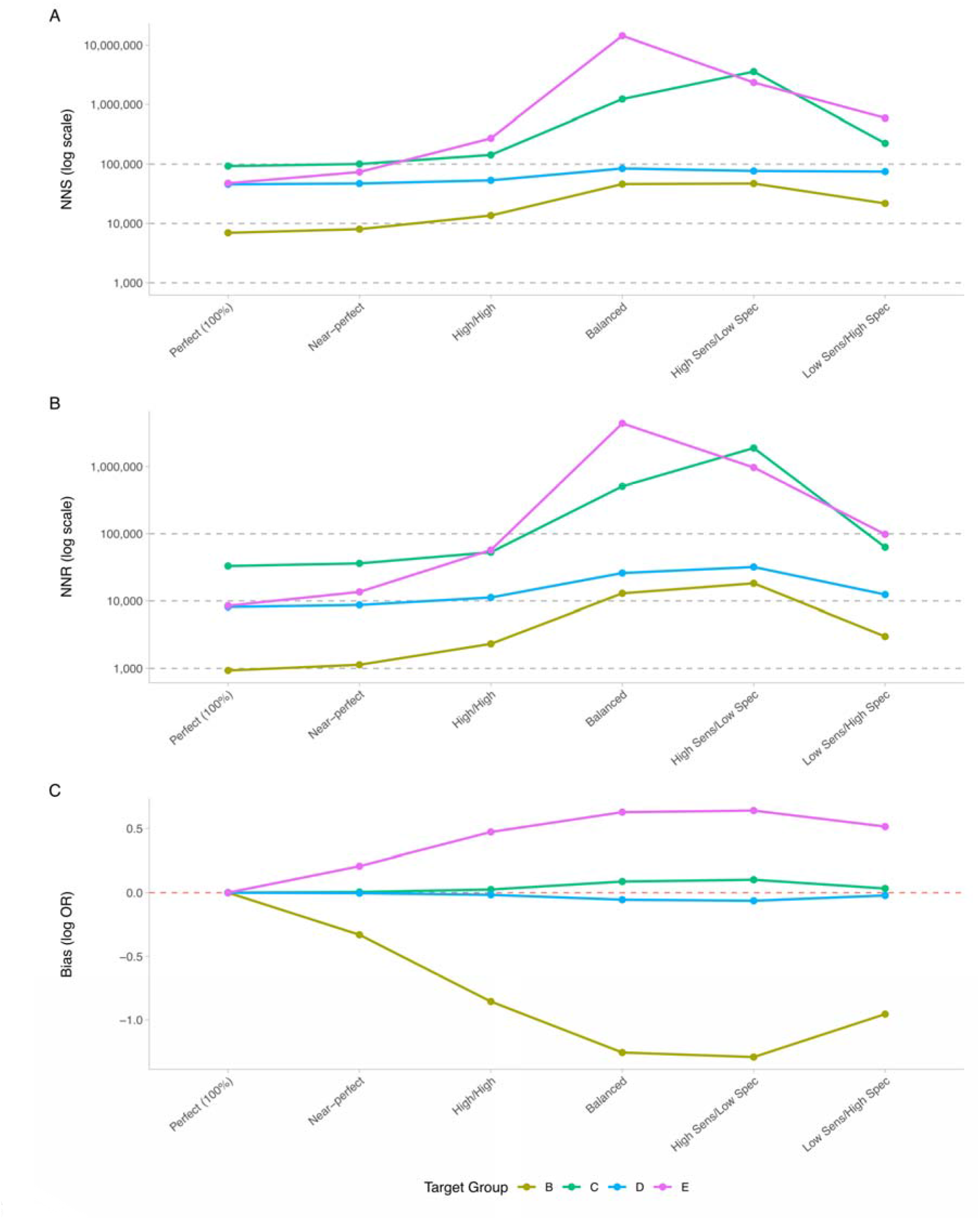
Number needed to screen (NNS), number needed to randomise (NNR), and bias across the four non-null subgroups. The x-axis shows test-performance scenarios; y-axes show NNS, NNR, or bias. Dashed horizontal reference lines indicate 1,000, 10,000, and 100,000 participants.

### Ordinal outcomes can substantially improve power, but the trade-off depends on the outcome scale and treatment-effect mechanism

Finally, we tested whether ordinal outcomes could rescue power lost by binary analyses in this setting. We simulated a 6-point ordinal outcome using the same subgroup setup as in ARREST under two scenarios: (1) a proportional-odds shift across the outcome scale, and (2) a non-proportional death-only effect (treatment changes death odds, with non-death categories rescaled proportionally). Note, the labels on this just correspond to potential outcomes, and simply are ordered by preference (e.g. death worst). The corresponding category shifts for subgroup B are shown in **Figure 4**. It should be noted that in ARREST, rifampicin was associated with worse outcomes in subgroup B, so the modelled effect is negative.

**Figure 4:**
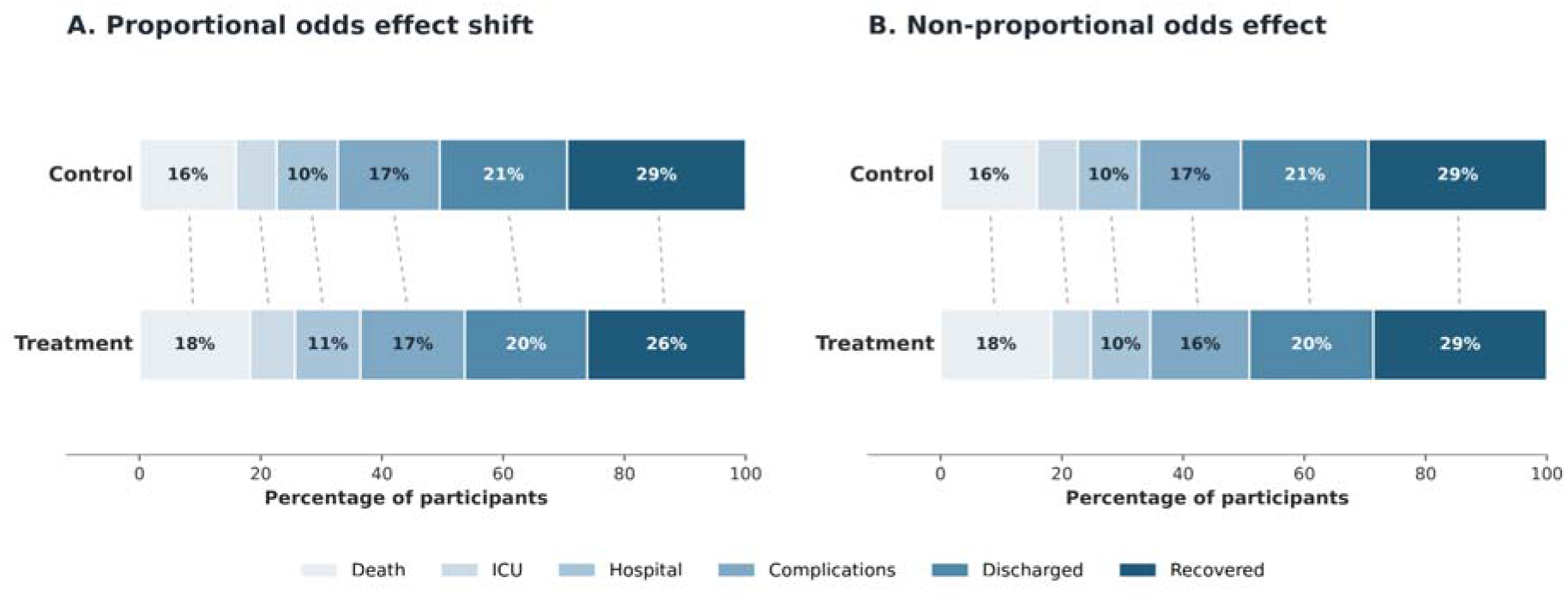
Ordinal outcome category shifts under proportional-odds (A) and non-proportional (death-only) effects (B) for subgroup B. Stacked bars show the control and treatment distributions across the six ordered categories (death, ICU/ventilated, still hospitalised, discharged to rehab, discharged with complications, discharged well). Within the non-proportional panel, survivor categories retain their relative composition and change only through rescaling after the treatment-associated shift in death. Note that this ARREST-derived subgroup B scenario models a harmful treatment effect at 84-day mortality.

When the ordinal outcome was appropriate, the ordinal model substantially increased power while maintaining Type I error control (**Figure 5**; **Supplementary Tables S4–S8**). Unexpectedly, under misclassification, ordinal estimates also tended to show lower absolute bias than binary estimates. This finding is explored in the discussion, but reflects the non-collapsibility^14^ of odds ratios which affects binary analyses more than ordinal ones.

**Figure 5:**
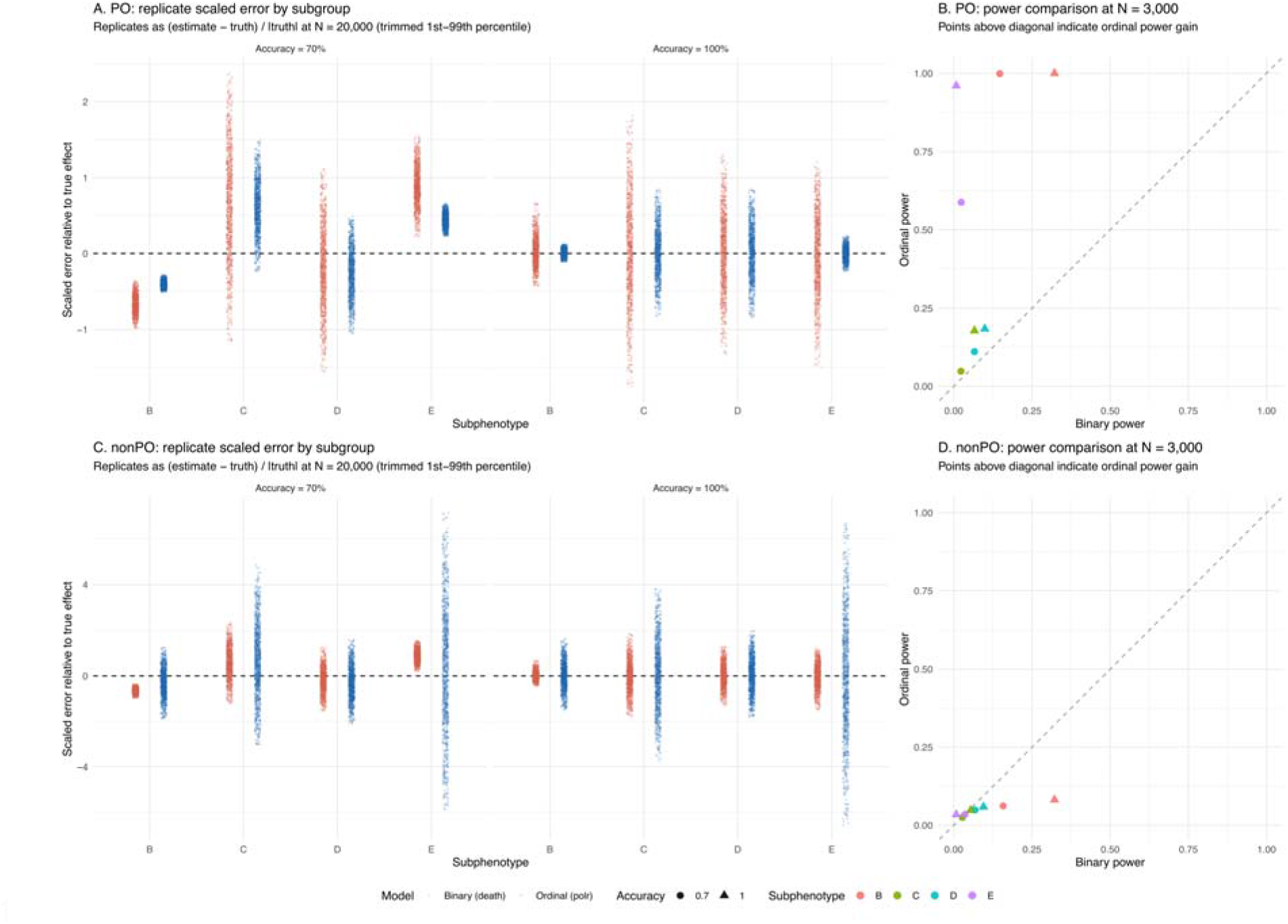
Binary versus ordinal performance under proportional-odds and non-proportional (death-only) ordinal data-generating mechanisms, stratified by classification accuracy. Panels A and C show replicate scaled error at n=20,000; panels B and D show power comparisons at n=3,000.

At n=3,000 and 100% accuracy, under proportional odds the ordinal model was more powerful than binary in all non-null subgroups: C (0.183 vs 0.109), D (0.214 vs 0.123), and E (0.943 vs 0.108), although B was 100% powered for both. Under non-proportional death-only effects, binary death analysis was more powerful for B (0.999 vs 0.913), C (0.102 vs 0.081), D (0.134 vs 0.090), and E (0.108 vs 0.045), although the reduction in power was less prominent.

Importantly, the degree of ordinal underperformance depends on how the ordinal scale is constructed. Additional ordinal performance and calibration detail are provided in Supplementary Tables S4, S5, S6, S7, and S8. A 5-point versus 6-point ordinal-scale sensitivity analysis is presented in Supplementary Table S9; with little compelling difference.

## Discussion

This simulation study shows that detecting heterogeneous treatment effects in SAB subgroups is more challenging than the headline estimates in the current literature suggest. Feasibility is determined not simply by the magnitude of a putative effect, but by the interplay of subgroup prevalence, baseline event rate, classification accuracy, and outcome choice.

### Sample size requirements and the challenge of planning

Even under perfect classification, required sample sizes remained large for realistic effect sizes for previously described subgroups. Subgroup B, however, was clearly detectable at realistic sample sizes, whereas C and D remained difficult and E remained strongly dependent on outcome choice. Critically, subgroup prevalence and mortality varied substantially across the published cohorts we examined, producing marked variation in required sample size for the same subgroup. This makes a priori sample size estimation for subgroup-specific effects unreliable, as the distribution of subgroups in a planned trial is unlikely to match that of any single historical cohort. Adaptive designs, such as Bayesian platform approaches with information-borrowing, may therefore be preferable to fixed designs predicated on a single assumed prevalence.

### Misclassification: power loss, bias, and the limits of enrichment

Imperfect classification compounded these challenges (even assuming that each subgroup represent a stable latent trait, which is unlikely^16^). Even modest reductions in accuracy produced substantial power loss and, more importantly, biased effect estimates. This bias arises because misclassification mixes patients from subgroups with different true effects, pulling observed estimates towards the population average. The direction and magnitude of this dilution depends on which subgroups patients are misclassified into, making the resulting bias difficult to predict or correct analytically. For rare subgroups with large effects - precisely the subgroups of greatest clinical interest - even small misclassification rates severely attenuated the observed effect. In the worst case, misclassification can produce estimates in the wrong direction. This is arguably a greater concern than simple power loss.

Enrichment strategies, in which only patients classified into a target subgroup are randomised, reduced the number needed to randomise but did not resolve bias and imposed screening burdens that remained substantial for most subgroups. Subgroup B, with its combination of large effect and moderate prevalence, remained a plausible enrichment target. For the remaining subgroups, even near-perfect classifiers produced screening requirements that were still difficult to justify.

Enrichment therefore looks useful in selected settings rather than as a general solution to underpowered subgroup analyses.

### Ordinal outcomes: power, bias, and the non-collapsibility mechanism

Our finding that ordinal outcomes can reduce bias arising from subgroup misclassification, alongside their established power advantages, has not previously been quantified in this setting to our knowledge. The mechanism derives from non-collapsibility.^14^ The odds ratio is a non-collapsible effect measure: mixing groups with different baseline risks causes the marginal estimate to depart from the true conditional effect, even in the absence of confounding. This departure is most severe when event probabilities are extreme. Binary mortality endpoints concentrate estimation at a single threshold where, for low-mortality subgroups, non-collapsibility is maximal. The proportional odds model estimates the same odds ratio but pools information across multiple cumulative thresholds, some with less extreme probabilities, and is consequently less affected. This advantage was most pronounced for rare, low-mortality subgroups—precisely those where binary analyses are most vulnerable.

Crucially, this advantage only holds when the proportional-odds assumption is approximately correct. When the treatment effect was concentrated mainly at death (our death-only simulation), the ordinal model diluted part of the signal with uninformative thresholds and binary analysis remained superior. The magnitude of this penalty was not fixed, however, and depended in part on the number of outcome levels used: adding levels improved efficiency under proportional-odds scenarios but could reduce robustness under death-only non-proportional scenarios. This has direct implications for outcome selection. This effect on death only has previously been demonstrated in infection trials.^17^

### The case for mechanism-matched outcomes in infection trials

Our results contribute to an ongoing debate about outcome standardisation in infection trials that extends well beyond SAB. There is a compelling case for core outcome sets, as promoted by COMET^18^ and widely used in recent trials.

Standardisation facilitates meta-analysis, reduces selective reporting, and enables cross-trial comparison. In SAB, where trials have variously used 14-day, 30-day, 84-day, and 90-day mortality with or without microbiological or complication-based composites,^3^ the appeal of a single agreed outcome is obvious.

However, our simulations demonstrate a fundamental tension with this approach. The optimal outcome depends not only on what matters to patients and clinicians, but on the mechanism by which the intervention is expected to act, which is likely to differ across different interventions. By this latter measure, a single core outcome cannot simultaneously be optimal for all interventions for the same disease.

Several recent infection trials illustrate this. In CAMERA2^19^, the addition of flucloxacillin to vancomycin for MRSA bacteraemia appeared to improve microbiological clearance while possibly worsening clinical outcomes. An ordinal outcome incorporating microbiological endpoints would have been well powered to detect an overall "effect," but that effect would have been in the opposite direction to the clinically important harm signal. Conversely, the success of the WHO ordinal scale in COVID-19 trials^20^ likely reflects the fact that immunomodulatory interventions such as dexamethasone had broad effects across the severity spectrum, making the proportional odds assumption approximately correct. Had those trials studied a narrowly acting intervention, the ordinal scale would have been less efficient than a targeted binary endpoint, precisely as our simulations predict.

The implications for SAB are substantial. Different interventions under consideration plausibly act through different mechanisms. Adjunctive antibiotics such as rifampicin or fosfomycin are hypothesised to improve bacterial clearance, which might manifest across the outcome spectrum - fewer complications, shorter hospital stays, and reduced mortality - making ordinal outcomes well suited. Source control interventions such as surgery for endocarditis act on a specific anatomical problem, where the relevant outcome may be prevention of a defined complication rather than a graded improvement. Host-directed therapies might primarily affect the most severely ill, with effects concentrated at the mortality threshold. When subgroups are added to this picture, the complexity increases further: the same intervention might plausibly act differently across subgroups. In our simulated subgroup B, despite only modest baseline mortality, the clinically relevant endpoint might still include complications or trajectory rather than death alone, whereas in higher-severity subgroups mortality may be the most informative measure. A well-designed platform trial in SAB might therefore need to pre-specify different primary outcomes for different intervention-subgroup combinations - an approach that is methodologically coherent but presents obvious challenges for interpretation and regulatory acceptance.

We do not think this complexity should discourage core outcome sets for SAB. Standardised outcomes remain essential for comparability and should be reported in all trials.^3^ However, our data suggest they should function as a minimum reporting standard rather than as a constraint on the primary endpoint. Trialists should be encouraged to select a primary outcome matched to the hypothesised mechanism while still reporting the core set for cross-trial comparison. This distinction between "always report" and "always analyse as primary" is important and is sometimes lost in advocacy for outcome standardisation. More broadly, the choice of outcome is itself a scientific hypothesis about how an intervention works and should be justified as such in trial protocols. Simulation studies tailored to the specific disease and intervention context can help quantify the consequences of outcome choice and should become a routine part of trial design.

### Comparison with previous literature

To our knowledge, no study has specifically quantified sample sizes, bias, and power for patient stratification in SAB, although the dependence of these quantities on effect sizes, event rates, and sample size is well established. The novelty here lies in deriving SAB-specific parameters from stratification across multiple clinical cohorts and a randomised trial. We note an important difference from McHugh et al., who showed ordinal outcomes were more powerful than binary outcomes across all settings.^21^ Our results differ because we fixed the binary comparison at death versus survival - where the treatment effect was concentrated in our non-proportional scenario - while they used a functional binary classification. This difference underscores how sensitive conclusions about outcome choice are to simulation assumptions. Finally, some recent literature has explored when outcomes should be measured in SAB, finding that early deaths were more likely related to SAB itself rather than any underlying comorbidity. Although not directly relevant, this further makes the point that outcome choice should be determined by the plausibility that the intervention affects the outcome. Outcomes that are not plausibly caused by treatment (or only partially) are less useful and less powered than specific outcomes.

## Limitations

The principal limitation of any simulation study is that its data-generating mechanisms may not reflect clinical reality. Whether binary analyses outperform ordinal analyses depends strongly on how outcomes are generated, and our results should therefore be interpreted primarily as illustrating the scale of the sample-size problem and the importance of outcome design. Our primary parameters derive from Swets et al., whose latent class analyses span multiple cohorts from the UK, Spain, the Netherlands, and the USA. ^8,9^ A recent independent study identifying a broadly similar set of subgroups provides some reassurance about their validity,^11^ but the current model still assigns every patient to a class; in reality, some patients will be difficult to classify, and future work should develop improved assignment models. In clinical syndromes such as SAB, where host response and disease severity are likely continuous, overlapping and temporally dynamic, hard class labels discard classification uncertainty and may cause bias in downstream subgroup analyses.

Our misclassification model also assumed random redistribution proportional to subgroup frequency, which is almost certainly optimistic. Structured misclassification between clinically similar subgroups, for example between the low-mortality subgroups B and E, could produce worse and less predictable bias patterns. Finally, the 50% winner’s-curse shrinkage applied to the subgroup effect estimates is arbitrary; if the true effects are smaller, the conclusions become correspondingly less optimistic.

## Conclusions and recommendations

Our results suggest a three-pronged strategy for advancing stratified trials in SAB. First, diagnostic classifiers must improve towards near-perfect performance if even moderately rare subgroups are to become viable trial targets. This is unlikely to be achieved using routinely available clinical data alone and is likely to require biomarkers of differential pathophysiologic processes and better understanding of the underlying biology. Second, trial resources should pragmatically focus on reproducible subgroups with favourable prevalence and event-rate characteristics, because attempts to power trials for rare, low-event subgroups with current methods are likely to fail. Third, and perhaps most importantly, outcome measurement should be matched to the plausible mechanism of the intervention under study. No single outcome will be optimal across all interventions and subgroups, and accepting this will require a shift in how infection trials are designed, reported, and interpreted.

## Ethics

No ethics was required for this study as purely a simulation study using publicly available published data.

## Patient Consent Statement

This simulation study used only publicly available, aggregate data from published studies and did not involve human participants, individual patient data, or identifiable information. Patient consent was therefore not applicable, and ethical approval was not required.

## Data and code availability

All code is available at https://github.com/gushamilton/sab_het

## Data Availability

All data is available at the linked github repo.

https://github.com/gushamilton/sab_het

