## Supplementary material for "Impact of subgroup classificuation accuracy on detecting heterogeneous treatment effects in *Staphylococcus aureus* bacteraemia: A simulation study": Sup methods

Supplementary Methods

We simulated individually randomised two-arm SAB trials to evaluate how subgroup misclassification affects estimation of subgroup-specific treatment effects.^1^

### Aims

The study evaluated how imperfect assignment of patients to clinically defined SAB subgroups affects the ability of a randomised trial to detect heterogeneous treatment effects.^2–5^

The primary aim was to quantify the effect of subgroup misclassification on power, Type I error, and bias when estimating subgroup-specific treatment effects on 84-day mortality. Secondary aims were to examine whether cohort composition changes the required sample size, whether enrichment designs could make subgroup-specific trials feasible, and whether ordinal outcomes could improve efficiency compared with binary death alone.

### Clinical inputs

Each replicate represented a two-arm, individually randomised trial with 1:1 allocation. The treatment contrast followed ARREST, a trial of adjunctive rifampicin versus placebo for SAB. Subgroup frequencies, baseline 84-day mortality, and subgroup-specific odds ratios were derived from ARREST subgroup analyses and related SAB subgroup studies. Because subgroup treatment effects identified after a trial are likely to be overestimated, the primary simulations used odds ratios defined by 50% shrinkage of the ARREST-derived log-odds ratios toward the null.^2,3,6^

The odds ratios are treatment effects on death. Values above 1 indicate higher mortality with treatment in that subgroup; values below 1 indicate lower mortality. Thus, this was not a simulation in which the intervention was assumed to benefit all subgroups. Subgroup A was treated as a null-effect subgroup and was used to assess Type I error. Subgroup B had the largest harmful effect after shrinkage, while E had the largest protective effect on the odds-ratio scale.

**Table S1. Clinical subgroup assumptions used for the primary simulations.**

| **Subgroup** | **Clinical label** | **Frequency (%)** | **Control death risk (%)** | **Shrunk OR** | **log(OR)** | **Treatment death risk (%)** |
| --- | --- | --- | --- | --- | --- | --- |
| A | Elderly/comorbid | 15.5 | 21.7 | 1.00 | 0.000 | 21.7 |
| B | Nosocomial IV | 13.4 | 7.5 | 4.34 | 1.468 | 26.0 |
| C | Metastatic | 35.6 | 19.2 | 0.89 | -0.117 | 17.5 |
| D | CKD | 17.8 | 18.2 | 1.19 | 0.174 | 20.9 |
| E | IDU | 17.8 | 2.9 | 0.56 | -0.580 | 1.6 |

The primary odds ratios used in the simulations are shown in Table S1. Treatment death risks were calculated from the control risk p₀ and subgroup odds ratio OR as p₁ = OR × p₀ / {1 − p₀ + OR × p₀}. Because odds ratios are non-collapsible, subgroup-specific odds ratios were estimated within each observed subgroup rather than interpreted as marginal population effects.^7^

Subgroups A-E were treated as mutually exclusive clinical classes. CKD denotes chronic kidney disease and IDU denotes injecting drug use; “Nosocomial IV” denotes the nosocomial/intravascular-line clinical subgroup label used in the source subgroup classification.

### Data generating mechanism

#### Patient generation and treatment assignment

Each simulated patient record had four quantities: true subgroup S, treatment Z, observed subgroup S*, and outcome Y. S took values A-E. Z was 0 for control and 1 for treatment. Outcomes were generated from the true subgroup S. The fitted subgroup analyses used the observed subgroup S*. If S* differed from S, patients were analysed in the wrong subgroup, which is how classifier error reduced power and introduced bias.

For the binary mortality simulations, outcomes were generated from the true subgroup S. The control death risk in subgroup s was p₀(s). The treated death risk p₁(s) was calculated from β(s), the subgroup log odds ratio:

$$p₁(s) = \frac{exp\{\beta(s)\} \times p₀(s)}{1 - p₀(s) + exp\{\beta(s)\} \times p₀(s)}$$

Death was then sampled as a Bernoulli outcome with probability p₀(s) in the control arm and p₁(s) in the treatment arm.

The primary sample-size grid used total trial sizes of 500, 1,000, 1,500, 2,000, 3,000, 5,000, 10,000, and 20,000. The expected numbers in subgroups A-E at n=5,000 were approximately 775, 670, 1,780, 890, and 890; at n=20,000 they were approximately 3,100, 2,680, 7,120, 3,560, and 3,560. These expected subgroup counts help explain why the same treatment effect can be detectable in one subgroup but not another.

#### Subgroup misclassification

Subgroup classification was applied after true subgroup assignment. The accuracy parameter a was the probability that the observed subgroup label equalled the true subgroup label, and the same value of a was used for every true subgroup:

$$Pr(S* = s | S = s) = a$$

$$Pr(S* = t | S = s) = \frac{(1 - a) \times\pi(t)}{1 - \pi(s)}, t \neq s$$

Here π(t) is the population frequency of subgroup t. Wrong labels were therefore redistributed in proportion to how common the other subgroups were.

The primary misclassification grid used classification accuracies from 70% to 100% in five-percentage-point increments. The 100% setting represents perfect subgroup ascertainment. The 70% setting represents a deliberately challenging classifier in which nearly one-third of subgroup labels are wrong. Because errors were redistributed according to subgroup frequencies, observed subgroup prevalences changed slightly when a < 1. Performance was judged against the named true subgroup effect rather than the diluted effect among all patients carrying an observed label, because the question was how classifier error affects recovery of the intended clinical subgroup effect.

#### Cohort composition

To examine whether sample-size requirements were stable across plausible SAB trial populations, the subgroup prevalence and mortality inputs were varied using five cohorts: ARREST, Edinburgh, Edinburgh 2, IDISA, and SABG-PCS, derived from the original and follow up publications. Treatment effects were held constant across cohorts so that the analysis isolated the influence of cohort composition and baseline risk. Required sample sizes were calculated for 80% power at two-sided alpha 0.05 using the normal approximation to the log odds ratio with equal treatment allocation. A fixed-n simulation at n=5,000 was then used to show the operating characteristics under each cohort mix.

**Table S Methods 2. Cohort prevalence and mortality inputs. Values are percentages for subgroups A-E.**

| **Cohort** | **N** | **Subgroup prevalence A-E (%)** | **Mortality A-E (%)** |
| --- | --- | --- | --- |
| ARREST | 758 | 15.2, 14.1, 36.4, 16.0, 18.3 | 21.7, 7.5, 19.2, 18.2, 2.9 |
| Edinburgh | 458 | 32.1, 28.8, 22.5, 8.5, 8.1 | 43.8, 15.5, 24.3, 26.2, 5.3 |
| Edinburgh 2 | 463 | 29.6, 26.6, 25.9, 10.6, 7.3 | 48.0, 21.0, 14.0, 38.0, 4.0 |
| IDISA | 490 | 23.3, 23.1, 19.8, 17.6, 16.3 | 78.0, 39.0, 46.0, 43.0, 11.0 |
| SABG-PCS | 755 | 32.3, 31.5, 16.4, 10.6, 9.1 | 78.0, 20.0, 22.0, 31.0, 21.0 |

#### Enrichment designs

The enrichment simulations asked a different question: if a future trial randomised only patients predicted to belong to a target subgroup, how many patients would need to be screened and randomised? Each non-null subgroup, B-E, was considered as the target subgroup. Six diagnostic-performance scenarios were considered: perfect sensitivity/specificity, near-perfect 99%/99%, high/high 95%/95%, balanced 80%/80%, high-sensitivity/low-specificity 95%/70%, and low-sensitivity/high-specificity 70%/95%.

For a target subgroup, the enrolled population contained true-positive target patients and false-positive non-target patients. The true-positive rate was sensitivity times the target subgroup prevalence. The false-positive rate was (1 - specificity) times the combined prevalence of the non-target subgroups, with false positives distributed across non-target subgroups in proportion to their prevalences. Control and treated event risks in the enriched trial were weighted averages across this enrolled mixture. Required randomised sample size was calculated from the resulting diluted log odds ratio using the same 80% power, two-sided alpha 0.05 normal approximation. Number needed to screen was required randomised N divided by the expected enrolment fraction.

#### Ordinal outcome construction

The main ordinal analysis used a six-level ordered outcome, from worst to best: death, ICU/ventilated, still hospitalised, discharged to rehabilitation, discharged with complications, and discharged well.^8^ The numbers 1-6 were ordered outcome scores, not equally spaced measurements. The analysis used the ordering of the categories, not a numerical distance between adjacent categories.

Death probability was anchored to the subgroup-specific 84-day mortality risk. Among survivors, the remaining probability mass was split across the five non-death levels in fixed proportions: 8% ICU/ventilated, 12% still hospitalised, 20% discharged to rehabilitation, 25% discharged with complications, and 35% discharged well. The resulting control-arm category probabilities varied by subgroup because death risk varied by subgroup, but the relative distribution among survivors was held constant.

The five cumulative ordinal contrasts were: death versus all better outcomes; death or ICU/ventilated versus all better outcomes; death, ICU/ventilated, or still hospitalised versus all better outcomes; those categories plus discharged to rehabilitation versus all better outcomes; and all categories except discharged well versus discharged well.

**Table S Methods 3. Control-arm six-level ordinal outcome probabilities by subgroup (%).**

| Group | **Death** | **ICU/ventilated** | **Still hospitalised** | **Discharged to rehab** | **Discharged with complications** | **Discharged well** |
| --- | --- | --- | --- | --- | --- | --- |
| A | 21.7 | 6.3 | 9.4 | 15.7 | 19.6 | 27.4 |
| B | 7.5 | 7.4 | 11.1 | 18.5 | 23.1 | 32.4 |
| C | 19.2 | 6.5 | 9.7 | 16.2 | 20.2 | 28.3 |
| D | 18.2 | 6.5 | 9.8 | 16.4 | 20.5 | 28.6 |
| E | 2.9 | 7.8 | 11.7 | 19.4 | 24.3 | 34.0 |

Under the proportional-odds outcome model, outcomes were generated using a cumulative-logit model:

$$logit\{Pr(Y \leq k | Z, S = s)\} = \theta_{k}(s) + \beta(s)Z, k = 1,...,5$$

Y was coded from 1 (death) to 6 (discharged well), Z was 1 for treatment and 0 for control, and β(s) was the subgroup treatment effect. The baseline cut-points θₖ(s) were calculated by taking the logit of the control-arm cumulative probabilities in Table S Methods 3. Proportional odds means that the same β(s) was applied to every cumulative split. Because lower scores are worse, β(s) > 0 shifts probability toward worse outcomes and β(s) < 0 shifts probability toward better outcomes.

Under the death-only non-proportional outcome model, treatment changed only the probability of death. The treated death probability was calculated from the subgroup death odds ratio. Among survivors, the probabilities for levels 2-6 were rescaled so that their relative split stayed the same as in the control arm. This means treatment changed death but did not change the pattern of non-death outcomes among survivors.

### Fitted Models and Targets

Binary death analysis:

Within each observed subgroup S* = s, death was analysed using a logistic regression with treatment as the only predictor:

$$logit\{Pr(death | Z, S* = s)\} = \alpha(s) + \gamma(s)Z$$

The fitted treatment effect was γ(s). With perfect subgroup classification, the target value was β(s) = log{OR(s)} from Table S1.

Observed subgroups when classification was imperfect. When a < 1, patients analysed under S* = s were a mixture of true subgroups. The fitted model still used the observed subgroup label, because that is what would be available in a trial using an imperfect classifier. Bias and power were calculated against the original effect β(s) for the named true subgroup. For subgroup A, β(A) = 0; under imperfect classification, significant estimates in observed subgroup A can arise either from ordinary false positives or from contamination by non-null subgroups, so the reported value is best read as the false-positive rate for the intended null subgroup analysis.

Ordinal proportional-odds analysis:

Within each observed subgroup S* = s, the six-level outcome was analysed using a cumulative-logit model:

$$logit\{Pr(Y \leq k | Z, S* = s)\} = \alpha_{k}(s) + \gamma(s)Z, k = 1,...,5$$

The fitted treatment effect was the single coefficient γ(s), shared across the five cumulative contrasts.

Ordinal outcomes generated under proportional odds. When the outcome was generated with the same cumulative-logit form, the target for the ordinal model was β(s) = log{OR(s)}. The collapsed binary death model was fitted to the same simulated patients and was also judged against the death log odds ratio β(s).

Ordinal outcomes generated under the death-only model. For the binary death model, the target remained the subgroup death log odds ratio β(s). For the ordinal proportional-odds model, the target was not β(s), because the fitted model assumes one treatment effect across all cut-points while the simulated treatment effect was present only at death. The target was the value of γ(s) that best matched the death-only category probabilities under a proportional-odds model. This value was calculated numerically from the control and treated category probabilities before the performance summaries were produced.

### Simulation Scenarios

The main binary mortality simulations used 2,000 independent replicates for each combination of total sample size and classification accuracy. For the main misclassification figure, total trial size was fixed at 20,000 and 1,000 replicates were used to give a clearer high-resolution view across classification accuracies. The ordinal proportional-odds and death-only non-proportional comparisons used 1,000 replicates, classification accuracies of 70% and 100%, and the same total sample-size grid. A separate simulation of 200,000 participants with perfect subgroup labels was used to check the treatment-effect values used for judging ordinal estimates, especially the value approached by the proportional-odds model when the simulated treatment effect acted only on death.

The primary two-sided significance threshold was alpha 0.05. A Bonferroni-style alpha 0.01 threshold was also calculated as a focused sensitivity analysis for multiple subgroup testing under perfect classification at total trial sizes of 5,000 and 10,000. Significance was assessed using Wald confidence intervals for the fitted log odds ratio: an estimate was significant when the two-sided confidence interval excluded zero. All reported power estimates count only statistically significant estimates in the correct treatment-effect direction, rather than simply counting any significant result.

Binary models were fitted by logistic regression. Ordinal models were fitted by proportional-odds logistic regression. Fits with too few observations, no treatment variation, no outcome variation, or model-fitting failure returned missing estimates and did not contribute to the numerator of power or error summaries.

### Performance Measures

Power was defined as the proportion of replicates in which the fitted treatment effect was statistically significant and had the same sign as the true subgroup effect. For subgroup A, where the true effect was null, the corresponding performance measure was Type I error: the proportion of replicates with a statistically significant treatment effect.

Type S error was the proportion of replicates with a statistically significant estimate in the wrong direction. Type M error was the mean exaggeration ratio among statistically significant estimates, calculated as the absolute estimated log odds ratio divided by the absolute target log odds ratio; it was not calculated for subgroup A because the target effect was zero. Bias was summarised as both signed bias and absolute bias on the log odds scale. For the ordinal non-proportional analyses, proportional bias and root-mean-square error were also reported because the fitted proportional-odds model is deliberately misspecified under the death-only mechanism.
